# Delivery Characterization of SPL84 Inhaled Antisense Oligonucleotide

**DOI:** 10.1101/2023.01.09.23284328

**Authors:** Efrat Ozeri-Galai, Lital Friedman, Ofra-Barchad-Avitzur, Matthew R Markovetz, William Boone, Kaitlyn R Rouillard, Chava D Stampfer, Yifat S Oren, David B Hill, Batsheva Kerem, Gili Hart

**Affiliations:** SpliSense Biohouse Labs, Haddasah Ein Kerem, Jerusalem, Israel; David Hill- Marsico Lung Institute, University of North Carolina at Chapel Hill, Chapel Hill, North Carolina; Department of Genetics, The Hebrew University, Jerusalem, Israel

## Abstract

The last years have shown enormous advancement in the therapeutic potential of RNA-related treatments, specifically for antisense oligonucleotide (ASO)-based drugs, leading to increased numbers of ASO regulatory approvals. In this study we focus on SPL84, an inhaled ASO-based drug, developed for the treatment of the pulmonary disease, Cystic Fibrosis (CF). Pulmonary drug delivery is challenging, due to a variety of biological, physical, chemical, and structural barriers, especially when aiming to target the cell nucleus. The efficient distribution of SPL84 in the lungs, penetration into the cells and nucleus, and stability are critical parameters that will impact drug efficacy in a clinical setting. In this study, we demonstrate the proper distribution and cell and nucleus penetration of SPL84 in mouse and monkey lungs. In vivo and in vitro studies confirmed the stability and mobility of our inhaled ASO drug through CF patient-derived mucus and in lung lysosomal extracts. Our results, supported by a promising pre-clinical pharmacological effect, emphasize the high potential of SPL84 as an effective drug for the treatment of CF patients.

In addition, successfully tackling the lung distribution of SPL84 and specific cell targeting offers huge opportunities for further development of SpliSense inhaled ASO-based drugs for unmet pulmonary diseases.

## Introduction

Antisense oligonucleotides (ASOs) are single-strand small synthetic nucleic acid molecules, able to bind specific sequences within target RNA molecules. ASOs are usually 16–25 bases long and are designed to hybridize with RNA through Watson-Crick base-pairing. ASOs can have different mode of actions, one of which is splicing modification. Splice switching ASOs consist of chemically modified nucleotides which ablate RNase H activity and allow interaction with nuclear pre-mRNA during the splicing process (1). ASOs can be designed to bind to 5’ or 3’ splice junctions, or to exonic splicing enhancer or silencer sites, blocking the RNA–RNA base-pairing or protein–RNA binding interactions that occur between components of the splicing machinery and the pre-mRNA. In doing so, they can modify splicing in various ways, such as promoting alternative use of exons, exon exclusion, or exon inclusion(2,3).

ASO-mediated therapies are clinically validated, in view of regulatory approvals of splice switching ASO-mediated therapies for the treatment of two rare diseases, Duchenne Muscular Dystrophy (DMD) and Spinal Muscular Atrophy (SMA) (4,5).

The key challenge for ASO-based therapeutics is the delivery of the ASO to its target site of action within tissues and to specific cells, while minimizing exposure of other tissues. Various strategies (such as chemical modification of the oligonucleotide itself, peptides or aptamers conjugates) are being pursued for the improvement of oligonucleotide effectiveness and specificity.

SPL84 is an inhaled ASO drug with the 2’-O-MOE phosphorothioate chemical modification developed for the treatment of Cystic Fibrosis (CF) patients carrying the 3849 +10kb C->T Cystic Fibrosis Transmembrane Conductance Regulator (CFTR) mutation. The 3849 +10kb C->T mutation is a splicing mutation leading to the inclusion of a cryptic exon that includes a premature termination codon (PTC). The PTC leads to degradation of a significant fraction of the mRNA by the nonsense mediated mRNA decay (NMD) mechanism as well as to the production of truncated non-functional CFTR proteins(6). SPL84 treatment results in splicing modulation, which leads to an increase of correctly spliced CFTR RNA and fully functional CFTR proteins(7).

A battery of in vitro pharmacological studies with SPL84 demonstrated its efficiency and potency, as reflected by full restortion of CFTR function in Human Nasal Epithelial (HNE) and Human Bronchial Epithelial (HBE) cells derived from CF patients carrying the 3849 + 10kb C->T mutation, measured using the Ussing Chamber assay(7). This assay serves as gold standard for efficacy assessment of CF drugs, due to the limitations and lack of CF animal models. The Ussing assay is considered as a strong predictor of patient response to the CF treatment (8–10).

As SPL84 is delivered to the lung via inhalation, bridging between the promising in vitro data of SPL84 in patient-derived airway cells to in vivo lung delivery and distribution was key in moving SPL84 into the clinic (Phase 1/2 study ongoing).

Lung architecture is commonly visualized as a bunch-of-grapes formed by 23 serial bifurcations from the trachea to the last alveolar duct, with 85–90% occupied by alveoli(11). The translocation of oligonucleotide-based drugs across the pulmonary mucosal epithelia is hampered by several biological barriers. These include the physical barriers of the overlying mucus layer and the tight packing of epithelial cells, combined with the sweeping movement of apical cilia that removes luminal material away from the mucosal surface.

In the present study, we confirmed the high stability of the chemically modified SPL84 in CF patient-originated mucus and in lung lysosomal extracts. We found that SPL84 properly distributes in mouse and monkey lungs following inhaled administration. Moreover, we demonstrated that SPL84 penetrates through the physical barrier formed by thick, diseased mucus, enters the different types of epithelial cells, and is released into the cell nuclei, which is the targeted compartment of the drug, both in vitro and in vivo.

## Material and Methods

### Synthesis of ASO

SPL84 is a uniformly modified 2-O-(2-methoxyethyl) phosphorothioate Antisense Oligonucleotide (ASO) composed of 19 nucleotide bases, in the form of a sodium salt. SPL84 was manufactured at Biospring. For the monkey study, SPL84 was manufactured at LGC, Biosearch Technologies Inc. Petaluma, CA, USA (FDA EI 3011897291). SPL84 was dissolved in 0.9% Sodium Chloride (pH 7.4, Baxter)/ PBS at the desired concentration for in vitro studies or animal administration.

### Animal experiments

#### Animal management

Mice experiments were performed in Science in Action, Ness Ziona, Israel. Animal handling was performed according to guidelines of the National Institute of Health (NIH) and the Association for Assessment and Accreditation of Laboratory Animal Care (AAALAC).

For the WT mice experiments, 14 CD-1 mice (all females) were used; standard age ∼ 6-8 weeks at the outset of the study. For the βENaC experiments, WT mice C57BL/6N littermates of βENaC mice O’Neal colony CB6088 and βENaC mice C57BL/6N Scnn1b-Tg O’Neal colony CB6088 were used. (Marsico lung institute, UNC Chapel hill, North Carolina).

The monkey experiments were performed in ITR Laboratories, Quebec, Canada. All animal procedures were conducted according to the Animal Care Committee (ACC) of ITR. All animals used on the study were cared for in accordance with the principles outlined in the current “Guide to the Care and Use of Experimental Animals” as published by the Canadian Council on Animal Care and the “Guide for the Care and Use of Laboratory Animals”, a NIH publication. *Mice experiments*

Mice were dosed intratracheally once or thrice (every other day) with the selected doses of SPL84 (mg/kg) and were divided into groups according to the dosing regime. For each group there were at least 3 mice treated under the same conditions. Phosphate buffered saline (PBS) /saline was used as a control.

#### Monkey experiments

Cynomolgus monkey (Macaca fascicularis) (Worldwide Primates Inc), males and females young adult at the onset of treatment, were used.

SPL84 was administered via inhalation by oronasal face mask exposure. Monkeys were dosed once a week for four weeks, at achieved dose levels of 2 and 25 mg/kg/week (two males/sex/dose). Three days following the final dose, animals were necropsied, and three sections of each side of the lung (left and right cranial lobes) were collected at each dose for in situ hybridization evaluation.

#### Intrathecal (IT) administration of SPL84 in mouse

For intrathecal administrations of SPL84, the mice were anesthetized with isoflurane and hung vertically by a rubber band which holds the upper incisors. The mouse tongue was gently pulled out using forceps and a volume of 50μL delivered directly into the trachea using a micropipette and the mouse was allowed to recover immediately.

### In Situ Hybridization (ISH) analysis

Four parallel sections were mounted onto two slides – two sections per slide. Parallel slides were processed for ISH so that one slide was hybridized to the probe and the second one was processed without the probe as a negative control.

Sections were hybridized to a digoxigenin (DIG) labeled 2’-O-methyl (2’-OMe) RNA probe and detected with Alkaline phosphatase (AP) labeled anti-Dig polyclonal Ab (Roche Diagnostics, Cat#11093274910). After hybridization, sections were lightly counterstained with Nuclear Fast Red. This analysis took place at Smart Assays Biotechnologies Ltd. (Ness Ziona, Israel)

### Stability of SPL84 in Rat lung lysosomal lysate (RLuLL)

SPL84 working solution was mixed on ice with RLuLL (XENOTECH. Cat#R1000.pl 1910156) in the presence of protease inhibitor cocktail (Calbiochemset III; EDTA free Cat#539134). Resulting solution was incubated at 37°C and aliquots were drawn at 5’, 30’, 3, 8 and 24 hours, transferred to a fresh tube and immediately frozen in liquid nitrogen (the final concentration of the SPL84 in the collection tubes is 5μM). As a control to evaluate RLuLL nuclease potency, a nuclease non-resistance double strand (ds) RNA molecule was treated under the same conditions. Evaluation of SPL84 stability was performed by Northern blot analysis. (This analysis was performed at Quark Pharmaceuticals, Ness Ziona, Israel)

### Northern blots and hybridization with 3’ labeled Digoxigenin LNA oligonucleotide probe specifically detecting SPL84

Samples were diluted with loading sample buffer boiled for 5 min at 95°C, and removed immediately to ice. Each sample were run onto denaturing gels. Equivalent amounts of SPL84 from working solution were similarly treated and loaded on each gel as a positive control for gel migration of a non-degraded ASO. Blotted membranes were hybridized with 3’ labeled Digoxigenin probe and detected with anti-DIG-HRP-Ab (Roche, anti-Digoxigenin-POD Fab fragment cat# 11 207 733 910). Membranes were exposed to an X-ray film and developed. (This analysis was performed at Quark Pharmaceuticals, Ness Ziona, Israel).

### Stability of SPL84 in CF sputum

Sputum from 10 (5 female, 5 male) individuals with CF acquired from the UNC Adult Pulmonary Clinic were pooled. Samples were mixed by trituration and then rotated for 24 hrs at 40°C (12). Stability assays were run at 1, 3, 6, 8, 24, and 48 hrs for pooled sputum at concentrations of 5 μM SPL84 with the addition of protease inhibitors (20X Protease Inhibitor Cocktail Set III, EDTA-Free (Calbichem, cat # 539134). At the specified time points, 50 μL aliquots were transferred to nuclease free Eppendorf tubes, and immediately frozen at -80°C until analysis. Evaluation of SPL84 stability was performed by Northern blot analysis.

### Migration of SPL84 in CF mucus

Mucus was prepared by pooling washings from >50 cell cultures from >10 CF donors. The samples were concentrated in dialysis tubing against spectra gel to reach 8% solids. 2 and 4% solids samples were prepared by dilution from 8% solids stock. Once mucus was prepared to the prescribed concentration, it was loaded into a capillary tube and topped with a matched concentration of mucus with Cy5-labeled SPL84 on a 384 well plate and read on a Tecan plate reader. Well intensity was measured every 15 minutes for 24 hours. Controls were performed by measuring the diffusion of FITC in buffer and at each mucus concentration. Additionally, the diffusion of Cy5-labelled SPL84 in buffer (PBS) was measured. 3-5 tubes were tested for each condition.

#### Statistical Analysis

Statistical analysis of the effective viscosity of SPL48 was performed using the students t-test. Results from SPL84 were compared to the effective viscosity of the molecule in water as well as to the effective viscosity of FITC-labelled dextran. To determine if the viscosity of SPL84 and FITC-labelled dextrans is correlated with concentration, we used a pairwise Pearson’s test.

### Human Broncihal epithelial (HBE) Primary cells amplification and differentiation

HBE cells were expanded by growing the cells in PneumaCult™-Ex Plus Medium (AMP medium). After expansion, cells were seeded on porous filters (Transwell, Corning) in air liquid interface culture (ALI). After cell differentiation in ALI conditions, 50μM Cy5-SPL84 was added from the apical side (on top of the mucus).

#### Localization and accumulation of the SPL84

48 hours from the last ASO addition the filters were fixed for analysis of localization and accumulation of the SPL84 using a confocal microscope. Vectashield mounting medium containing DAPI was added, and the filter was placed on a microscope slide. For image analysis, ImageJ software was used.

## Results

### Intratracheal administration of SPL84 results in proper SPL84 lung distribution in WT mice

SPL84 distribution in WT mouse lungs was analyzed following intratracheal administration at four doses: 0.2, 1, 2 mg/kg. The analysis was performed on fixed lung sections using in-situ hybridization (ISH) with a labeled “sense” probe, which hybridizes specifically to the SPL84 antisense sequence. At all tested doses, SPL84 was distributed across the entire lung, with a dose related increase in the staining intensity (Figure 1A). Higher magnitude images demonstrate the presence of SPL84 across the respiratory epithelium, including in epithelial cells in the bronchus and bronchioles, as wells as in the distal alveoli regions (Figure 1B).

**Figure 1.**
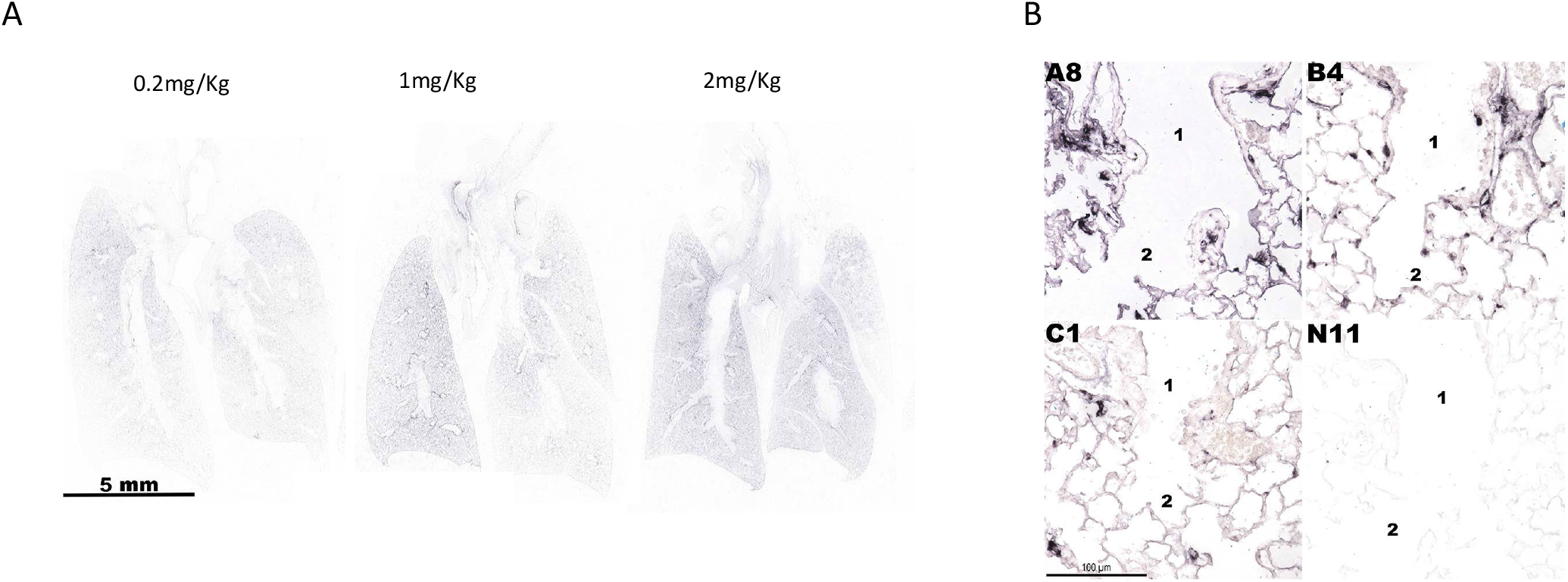
Proper distribution of SPL84 in WT mice lungs. ISH analysis for the detection of SPL84 in fixed mouse lung sections following IT administration at 4 different doses A. Low power scans of representative lung sections from all experimental groups B. High power images of representative lung samples from four experimental groups showing SPL84 localization detected by ISH. Images present respiratory epithelium at different airways levels and alveoli.(A8= 2mg/kg,B4=1mg/kg, C1=0.2mg/kg and N11=saline)

### Kinetics of lung distribution in WT mice following intratracheal dosing of SPL84

To assess the kinetics of lung distribution of SPL84 over 1 week in WT mice, mice were treated with a single dose or three (every other day) doses of 5 or 10 mg/kg SPL84 via intratracheal administration. Lungs were collected at 4 time points (1, 3, 5, or 7 days) after the final dose for each dosing regimen and fixed for ISH analysis. The results show that SPL84 was distributed and retained in the lung at all time points, up to 7 days post-dosing. Overall, there was no prominent difference in hybridization pattern and signal strength between the different groups; no difference was observed between the different dose levels (3 doses of 5 mg/kg vs 10 mg/kg), nor between the number of doses (3 doses vs 1 dose of 10 mg/kg) (Figure 2A). SPL84 was detected in cells in all areas of the lung, including the trachea, bronchi, bronchioles and alveoli (Supplementary Figure 1). A semi-quantitative evaluation of ISH signal in respiratory epithelium demonstrated that 3 days following the last administration, SPL84 was detected in >90% of epithelial surface cells, and 7 days post last administration, SPL84 was still widely spread along the epithelium (detected at 50-75% of the epithelial surface) (Figure 2B). Retention of SPL84 in the lungs 7 days following a single dose administration is supportive of weekly dosing in the clinic.

**Figure 2.**
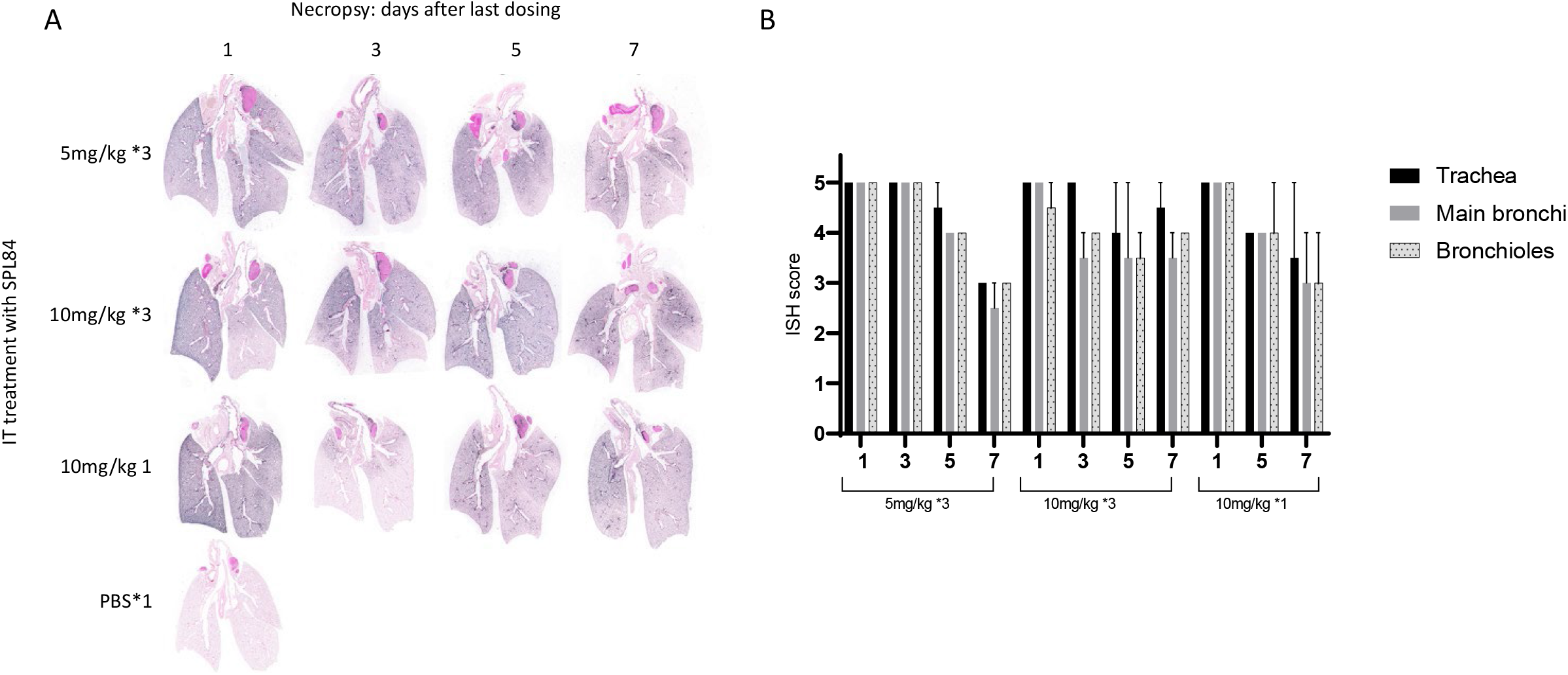
Kinetics of lung distribution of SPL84-treated WT mice for up to 1-week following intratracheal administration. A. Low power scans of representative lung sections from all experimental groups, treated as indicated with 1 or 3 IT administrations of SPL84 and collected at the indicated day following the last dosing. B. Semiquantitative analysis of the extent of hybridization signal throughout the respiratory epithelium. The approximate percentage of the labeled epithelium surface was evaluated and graded according to the following scale: Grade 0 – 0-10%, Grade 1 - 10-25%, Grade 2 - 25-50%, Grade 3 – 50-75%, Grade 4 - 75%-90%, Grade 5 >90%

We assessed the stability and retention of SPL84 in mouse lungs for a longer period by analyzing SPL84 distribution 4 weeks post treatment. Mice were treated with a single dose of 10 mg/kg SPL84 via intratracheal administration. Lungs were collected either 1, 2, or 4 weeks after dosing and fixed for ISH analysis. Results showed that SPL84 was distributed and retained in the lungs for up to 4 weeks post-dose. SPL84 was detected in respiratory epithelial cells at all parts of airways (Figure 3A). A semi-quantitative evaluation of ISH signal in respiratory epithelium demonstrated that a high coverage of the epithelium surface with SPL84 was maintained from 1 week up to 4 weeks following SPL84 administration (Figure 3B). Retention of SPL84 in the lungs for 4 weeks following a single dose is supportive of exploring less frequent dosing of SPL84 in the clinic, i.e., every two weeks, or less frequent, and these findings are aligned with the expected stability driven by the chemical modification of SPL84, as previously reported(13).

**Figure 3.**
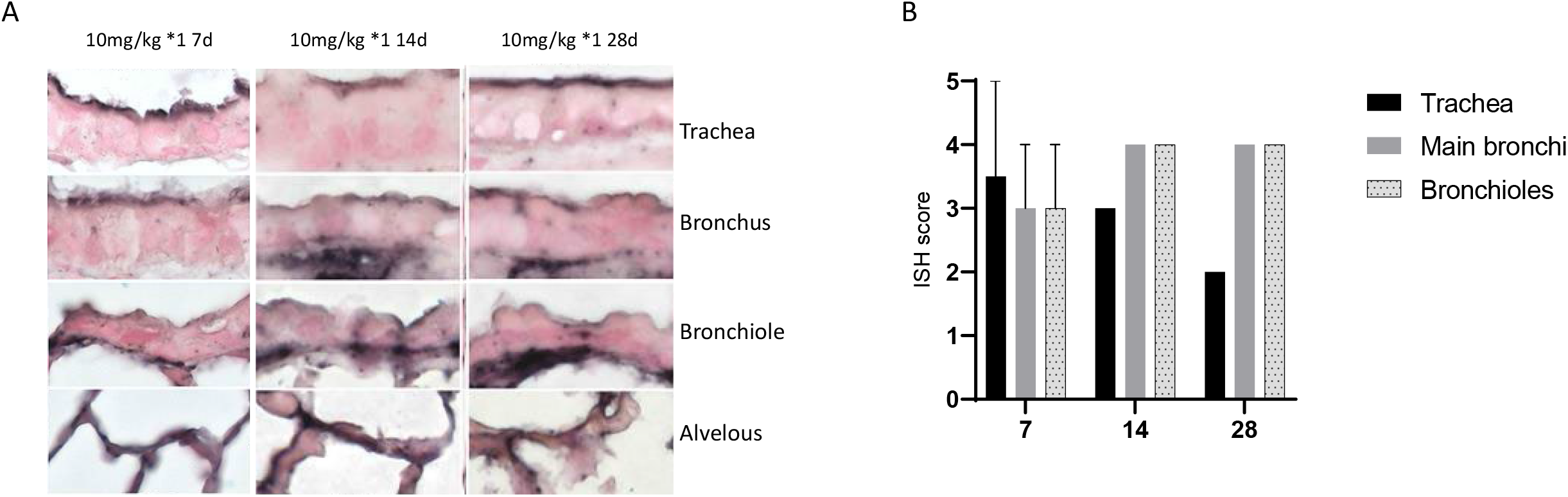
Kinetics of lung distribution of SPL84-treated WT mice for up to 4-weeks following an intratracheal administration A. High power microphotographs of representative lung samples from three experimental groups treated with a single dose of 10mg/kg, showing ASO localization detected by ISH. Lungs were collected at the indicated day following the treatment. Microphotographs present respiratory epithelium at different airways levels. B. Semiquantitative analysis of the extent of hybridization signal throughout the respiratory epithelium. The approximate percentage of the labeled epithelium surface was evaluated and graded according to the following scale: Grade 0 – 0-10%, Grade 1 - 10-25%, Grade 2 - 25-50%, Grade 3 – 50-75%, Grade 4 - 75%-90%, Grade 5 >90%

### Stability of SPL84 in rat lung lysosomal lysate

ASOs have been shown to enter cells through high-and low-binding plasma protein receptors on the cell surface, resulting in ASO compartmentalization into lysosomes and endosomes, as part of the natural process of endocytosis (14). A lysosome can break down many kinds of biomolecules, due to its composition of hydrolytic enzymes, combined with extreme acidic conditions. Therefore, we assessed the stability of SPL84 when spiked into rat lung lysosomal lysate (RLuLL). SPL84 was added to RLuLL and incubated at 37°C for up to 24 hours. Samples were taken over time and analyzed with northern blot using a SPL84 probe. Results showed that SPL84 incubated in RLuLL appear to be nuclease resistant and pH resistant for at least 24 hours, as indicated by the migration of the single oligonucleotide band to a position equivalent to that of the positive control sample (Figure 4A). In contrast, when a control nuclease non-resistant dsRNA was incubated in RLuLL, smaller migration bands, indicating degradation, appeared within 3 hours of incubation (Supp Fig 2). These results support the stability of SPL84 while entering the cells and into the nucleus.

**Figure 4.**
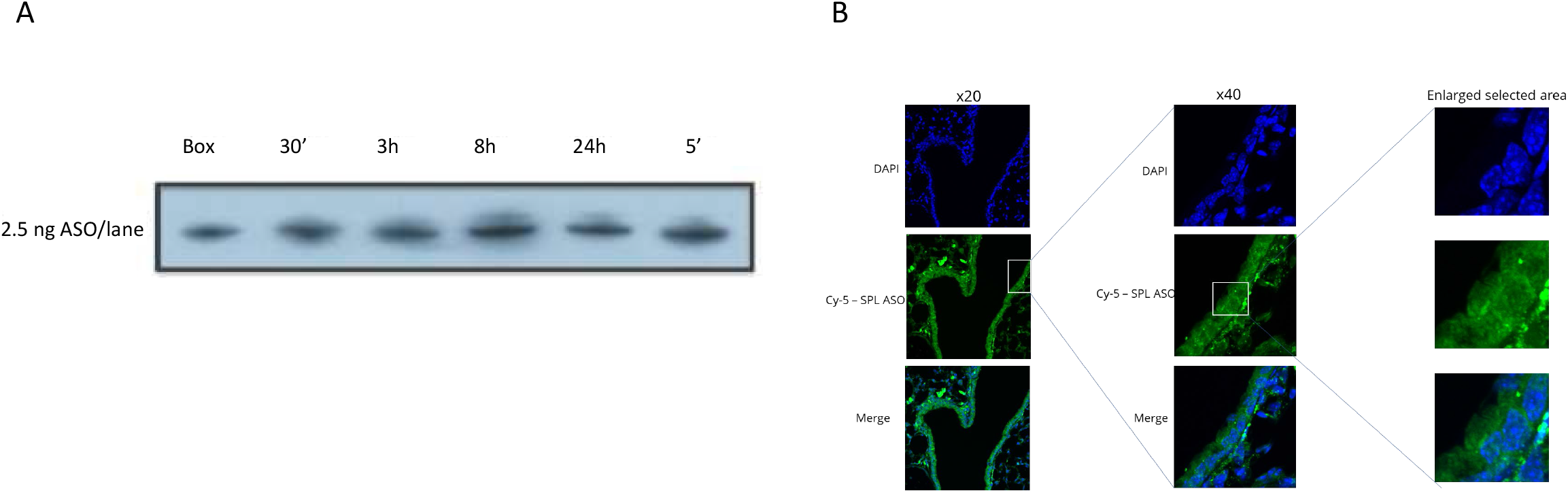
Stability of SPL84 and penetration to the epithelial cell nuclei. A. Northern blot of SPL84 samples incubated in rat lung lysosomal lysate (RluLL) for up to 24 hours. Box = positive control; untreated SPL84. ‘=min. B. An example of part of a mouse lung bronchus is presented. The presented confocal microscope image is a stack of only 4′,6-diamidino-2-phenylindole (DAPI) positive optical sections. Blue - nuclei (DAPI staining), Green - Cy5-SPL84.

### SPL84 enters lung epithelial cell nuclei in mice following intratracheal dosing

To directly evaluate the penetration of SPL84 to the nuclei of epithelial cells in-vivo, we assessed the lung distribution of a fluorescently labeled SPL84, allowing high sensitivity and resolution of detection. WT mice were dosed intratracheally 3 times (every other day) with Cy5-labeled SPL84. 48 hours following the last treatment, animals were sacrificed and necropsy was performed to collect trachea and lungs. Lung sections were processed, stained for nucleus detection and visualized using a confocal microscope. The presented images are a combination of the nucleus-containing optical sections only. As can be seen in Figure 4B, SPL84 penetrates efficiently into epithelial cells along the bronchus and is localized within the cell nuclei and cytoplasm. Similar results were obtained in distal (alveoli) regions (Supplementary figure 3). These results indicate that SPL84 is efficiently localized to the target region within the lung epithelial cells, even in distal regions.

### Proper lung distribution of inhaled SPL84 in monkey lungs

Monkeys are one of the large animal species frequently used for pre-clinical safety evaluation of drugs prior to clinical trials in humans. Monkeys bear close resemblance to humans in anatomical and physiological lung properties, providing a more adequate model system to study the lung distribution(15). Therefore, it is important to evaluate ASO distribution in the Monkey lungs.

Monkeys were treated with SPL84 once weekly via inhalation for 4 weeks at low and high doses (2 and 25 mg/kg/week). Three sections of each side of the lung were collected at necropsy, 72 hours after the last dose, and fixed for ISH analysis (Figure 5A).

**Figure 5.**
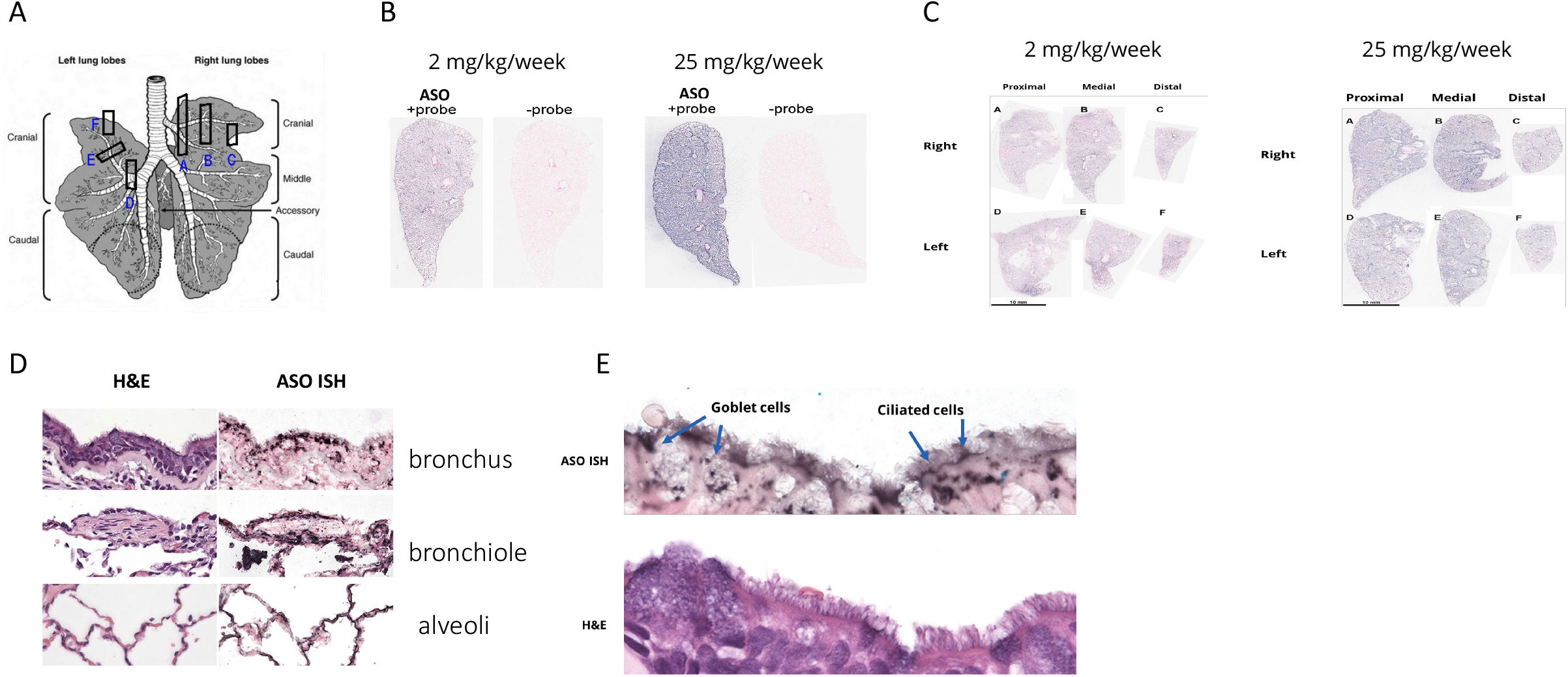
Proper lung distribution of inhaled SPL84 in monkey lungs. A. Schematic diagram of lung specimens collected for ISH study. B-C. Low power scans of and 25 mg/kg/week treated monkey lung sections hybridized to the SPL84-complementary probe. D-E. High power (objective x40) microphotographs of parallel hematoxylin-eosin stained (H&E) and hybridized (ISH) sections of bronchial epithelium from 25 mg/kg/week treated monkeys. Enlarge selected area (E).

We observed broad SPL84 distribution through the lungs, which was dose dependent (Figure 5B). The samples showed strong and uniform labeling in all sections sampled, from the proximal to the distal lung regions (Figure 5C). The SPL84 was detected across the respiratory epithelium (bronchi and bronchioles), as well as in the distal alveolar cells (Figure 5D).

A higher magnitude image also demonstrated that SPL84 enters different types of epithelial cells; ciliated and secretory cells (Figures 5E).

### SPL84 properly distributes through the mucus layer in a muco-obstructive mice model

Since CF is characterized by the accumulation of a thick mucus layer covering the airways, we investigated whether SPL84 penetrates the mucus layer in a muco-obstructive mouse model. The β-ENaC mice model was used for this study; these mice overexpress the epithelial Na(+) channel (ENaC) and generate a CF-like mucus in the mouse lung. WT and β-ENaC mice were treated with 1 and 5 mg/kg SPL84 via intratracheal administration every other day for 6 days. Lungs were collected 24 hours following the final treatment and fixed for ISH analysis. SPL84 was found to penetrate the mucus and properly distribute in the β-ENaC mice lungs. Comparable staining was observed in WT and β-ENaC mouse lungs, specifically in bronchial and bronchiolar epithelial cells at an estimated coverage of 90% (Figure 6).

**Figure 6.**
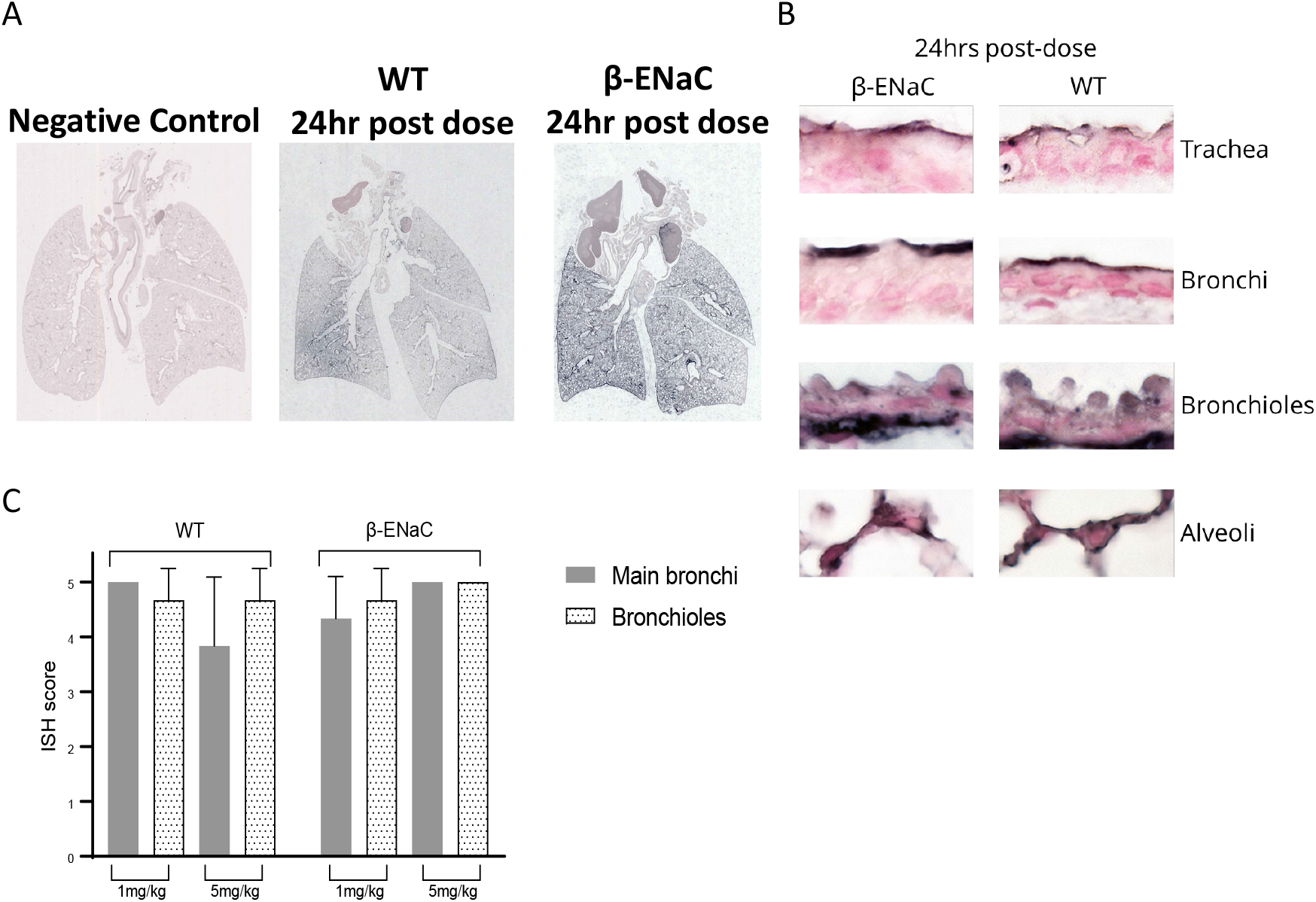
SPL84 properly distributes through the mucus layer in mouse model. A. Low resolution scans of sections from representative mice from different groups. From left to right: β-ENaC transgenic mice (TG) mice treated with saline, wild type (WT) mice treated with 5 mg/kg of SPL84, β-ENaC TG mice treated with 5 mg/kg of SPL84. B. High power β-ENaC TG (left) and WT (right) SPL84-treated mice at 5 mg/kg, hybridized to 2’-OME RNA probe. C. Semiquantitative analysis of the extent of hybridization signal throughout the respiratory epithelium in βENaC TG and WT treated SPL84-mice at 1 and 5 mg/kg. The approximate percentage of the labeled epithelium surface was evaluated and graded according to the following scale: Grade 0 – 0-10%, Grade 1 - 10-25%, Grade 2 - 25-50%, Grade 3 – 50-75%, Grade 4 - 75%-90%, Grade 5 >90%

### In vitro studies demonstrate stability, mobility, and penetration of SPL84 in CF sputum

The stability of SPL84 was further evaluated in the presence of sputum samples taken from 10 CF patients. SPL84 was incubated in CF sputum for up to 48 hours at 37°C in the presence of a protease inhibitor cocktail which protects nucleases against degradation. The analysis was done by northern blot using a SPL84 specific probe.

As shown in Figure 7A, the stability and integrity of SPL84 was maintained when spiked into CF sputum for up to 48 hours, as indicated from the single band in an equivalent position to that of the positive control sample. These results are aligned with the high stability of the 2’-O-MOE modified ASOs (16).

**Figure 7.**
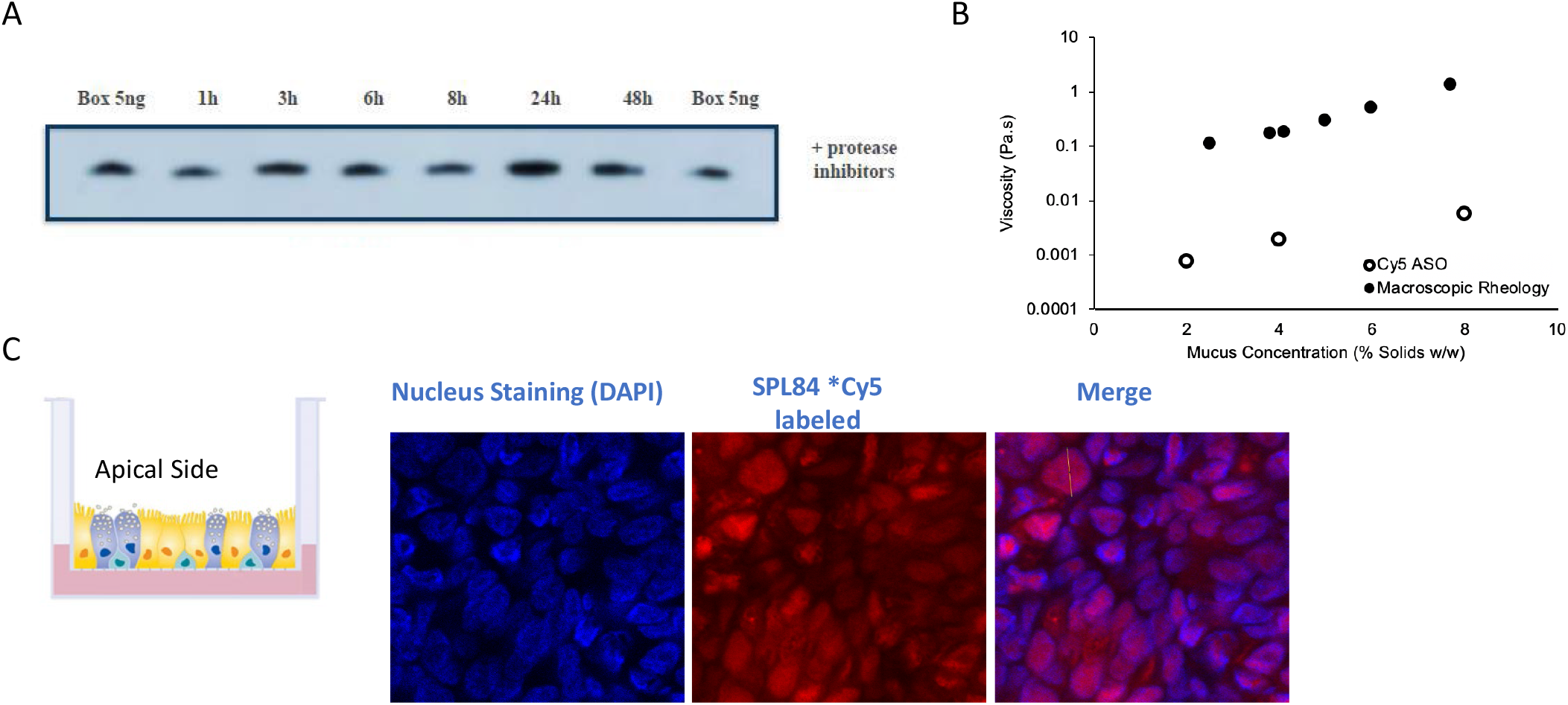
SPL84 demonstrates stability, mobility, and penetration of SPL84 in CF sputum. A. Autoradiograms of Northern blots following incubation of SPL84 (5μM) in mucus from CF patients at different time points, all samples were exposed to protease inhibitors. Box = positive control (SPL84 spiked in mucus from CF patients at 4°C was used for size control). B. Comparison of effective viscosity for SPL84 (signed in black circle, Cy5 ASO) compared to macroscopic rheology of mucus, as measured in a previous study (22). C. Schematic diagram of differentiated HBE cells seeded on transwell filter (left panel). HBE cells were differentiated for 35 days, the cells were allowed to accumulate mucus, which was not washed, and then treated with 3 additions of 50μM Cy5 labelled SPL84 every 2 days. 48hr post the last addition, the cells were fixed and visualized for detection of SPL84. The fluorescent signal was detected using a Cy5 filter under confocal microscope. Nuclei were stained with DAPI.

To test the ability of SPL84 to penetrate the human mucus layer, two in vitro studies were performed. In the first study, the migration of fluorescently labelled SPL84 in mucus harvested from an HBE cell culture was analyzed. The mucus was prepared to reach concentrations that mimic healthy (2% solids), mild CF (4% solids), and severe CF (8% solids) conditions, and SPL84 migration properties were compared to those of macroscopic particles. Although the effective viscosity for Cy5-labeled SPL84 slightly increased with mucus concentration, indicating a potential interaction with mucus, the SPL84 viscosity was ∼100-fold lower than macroscopic standard particles (Figure 7B). These data confirmed that while SPL84 interacts with mucus, these interactions are weak enough in nature and will not affect the penetration of even a pathological mucus layer by SPL84. Based on the measured diffusion coefficients, the maximum penetration time predicted in 8% mucus mimicking severe CF, is 85 seconds. In the second study, we assessed again the penetration of SPL84 through mucus generated in HBE cells grown in culture. HBE cells were grown in a 2D structure in air liquid interface (ALI). Upon cell differentiation, a pseudo-stratified epithelial layer is formed, consisting of a mixture of ciliated and mucus secreting epithelial cells, leading to the secretion of a mucus layer at the apical side of the cells. Following administration of fluorescently labeled SPL84 on the mucus layer at the apical side of the HBE cells, the SPL84 could be detected in the cell nuclei, confirming successful penetration through the mucus, cell penetration, and migration to the epithelial cell nuclei (Figure 7C).

Altogether, these results confirm that SPL84 can properly and efficiently penetrate and migrate through the mucus layer to epithelial cells and enter the nuclei where it modulates the CFTR pre mRNA in a specific manner, leading to the generation of full-length WT CFTR RNA.

## Discussion

Achieving optimal delivery and distribution of oligonucleotides to the lungs is in the scope of interest for many therapeutic approaches. We have developed SPL84, an inhaled ASO drug, targeted specifically for the treatment of CF patients carrying the 3849 +10kb C->T mutation. Administration of SPL84 via inhalation allows local administration to the target organ of the lungs, with minimal systemic exposure(17). One challenge of inhaled drug delivery for CF and other muco-obstructive diseases is the thick mucus layer covering the airways that can be an obstacle for drug delivery. The comprehensive results presented in this study demonstrate that SPL84 is properly distributed in WT and muco-obstructive mouse and WT cynomolgus monkey lungs. These results are in agreement with previous published data (17–19). Importantly, we demonstrated that inhaled SPL84 penetrates through the CF-like thick mucus layer into the epithelial cells both in vitro and in vivo. Moreover, since lungs are made up of many different cell types, it was important to show that SPL84 enters different types of epithelial cells - ciliated as well as secretory (goblet) cells. The proven durability of SPL84 for up to 4 weeks following a single dose in mice is supportive of a weekly / every other week inhalation regimen that will be further evaluated in clinical studies. This low frequency of administration will potentially provide a promising safety profile with reduced treatment burden on patients.

The small size (estimated as 1-3 nm), single strand, negatively charged SPL84, with demonstrated pronounced stability, confirmed that additional vehicles or components to support SPL84 delivery to the lungs is not required (14). On the contrary, it is proposed that larger, more complex inhaled RNA/DNA based therapies such as mRNA replacement, DNA editing or siRNA which require delivery systems based on encapsulated lipid nanoparticles (LNPs)/ specific ligand conjugates to allow penetration to lung epithelial cells or other cells might present some significant disadvantages. The physicochemical parameters of LNPs, including their size, shape, charge, and surface composition, directly impact cellular internalization efficiency and have also been associated with adverse immune events (20). Lastly, the larger size of these compounds could potentially result in significantly slower migration and penetration through the mesh generated by the thick disease mucus layer.

Taken together, the combination of functional rescue by SPL84 demonstrated in patient-derived respiratory cells with the proven efficient delivery to lung epithelial cells, along with the clinically validated chemical modifications (2’-O-MOE phosphorothioate, (13,21)) generates a promising drug that could potentially provide a cure for the lung disease of CF patients carrying the 3849 +10kb C->T mutation. In addition, there are additional, various severe pulmonary diseases, such as bronchiectasis, asthma, lung fibrosis, chronic obstructive pulmonary disease, and IPF, which can benefit from our established inhaled platform delivery system and ASO technology.

## Data Availability

All data produced in the present work are contained in the manuscript

## Declaration of interest and support

E.O, L.F, O.A, C.S, Y.O, and G.H are Splisense employees. B.K is the CSO of Splisense. This research is supported by Splisense LTD and by a Cystic Fibrosis Foundation TDA.

## Notes

### Author Declarations

Sputum samples were provided by the UNC Adult Pulmonary Clinic and collected under UNC IRB, # 15-2431. Mucus samples harvested from UNC cell cultures are acquired from the UNC Marsico Lung Institute Tissue Procurement and Cell Culture Core under UNC IRB protocol # 03-1396

