## Supplementary figures for "Delivery Characterization of SPL84 Inhaled Antisense Oligonucleotide"

### Supp Figure 1

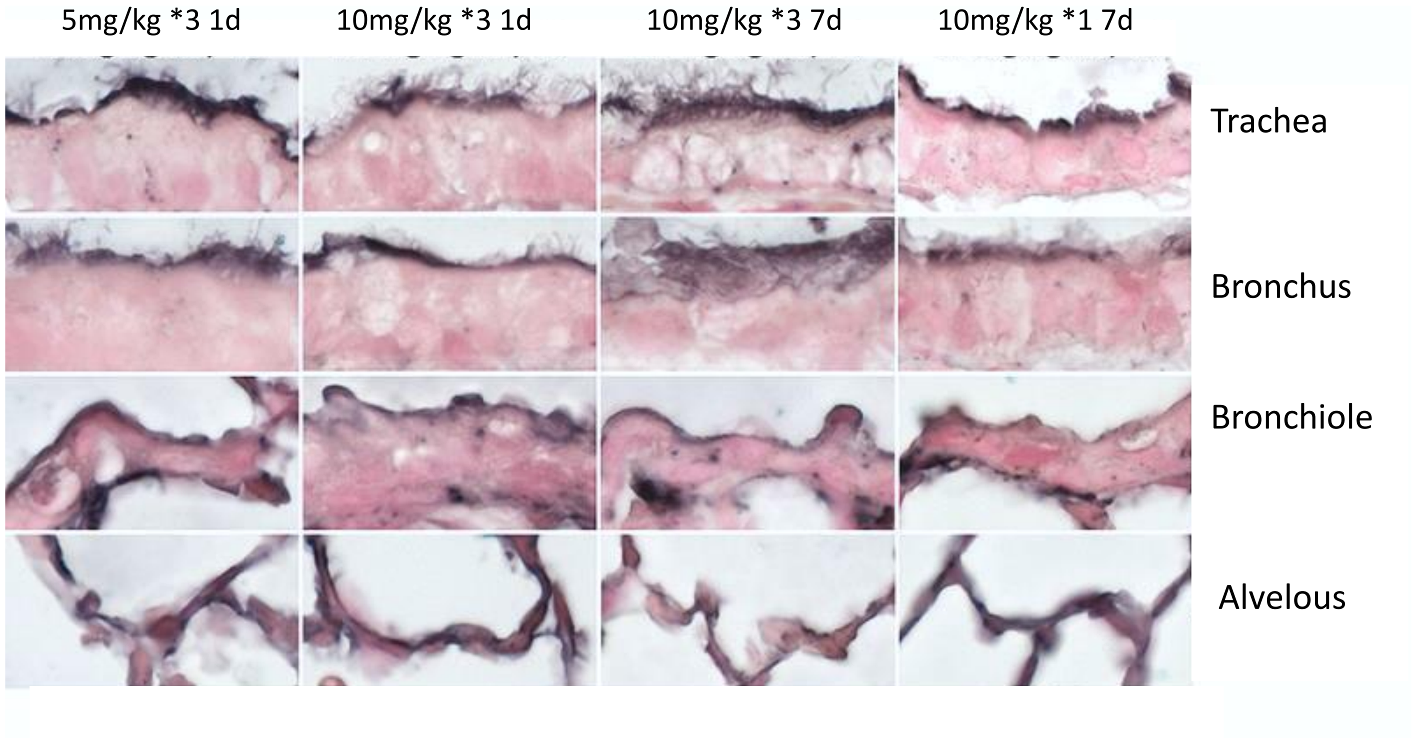


**Kinetics of lung distribution of the SPL84  WT mice for up to 4-weeks following intratracheal administration** A. High power microphotographs of representative lung samples from three experimental groups treated with three doses of 5mg/kg or 10mg/kg or single dose of 10mg/kg SPL84 showing ASO localization detected by ISH. Microphotographs present respiratory epithelium at different ay levels.

### Supp Figure 2

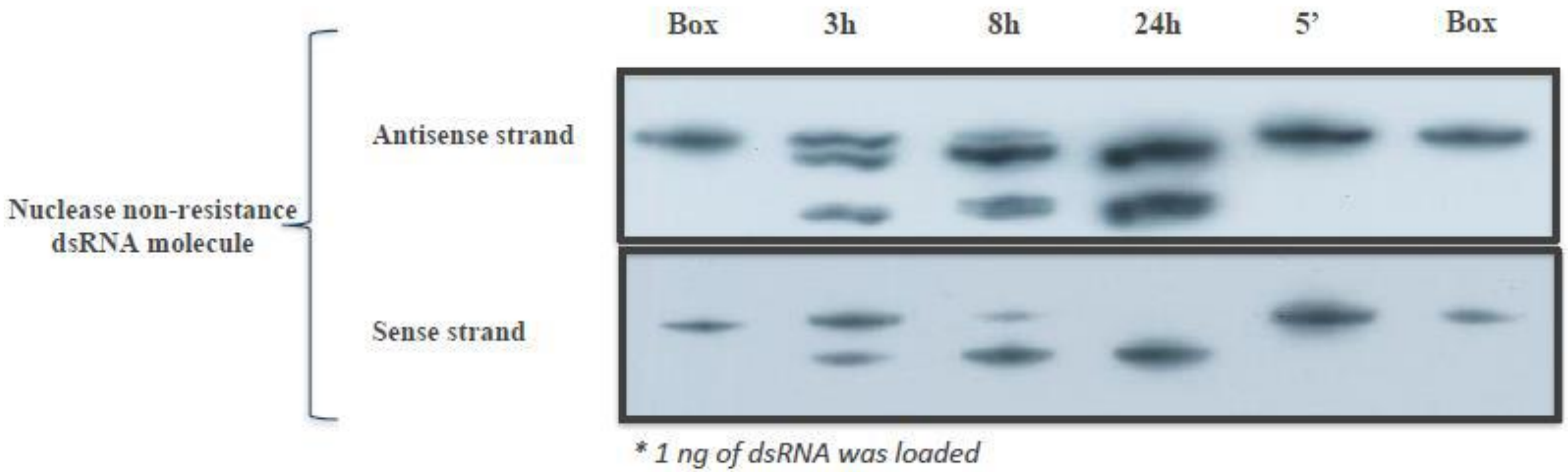


**Evaluation of Nuclease Potency in RLuLL Using a Control Nuclease Non-Resistant dsRNA.** Northern blot of nuclease non-resistant dsRNA samples incubated in RLuLL for up to 24 hours. Each of the membranes was hybridized with the relevant probe for either the sense or antisense strand. Box = untreated dsRNA. ’=min.

### Supp Figure 3

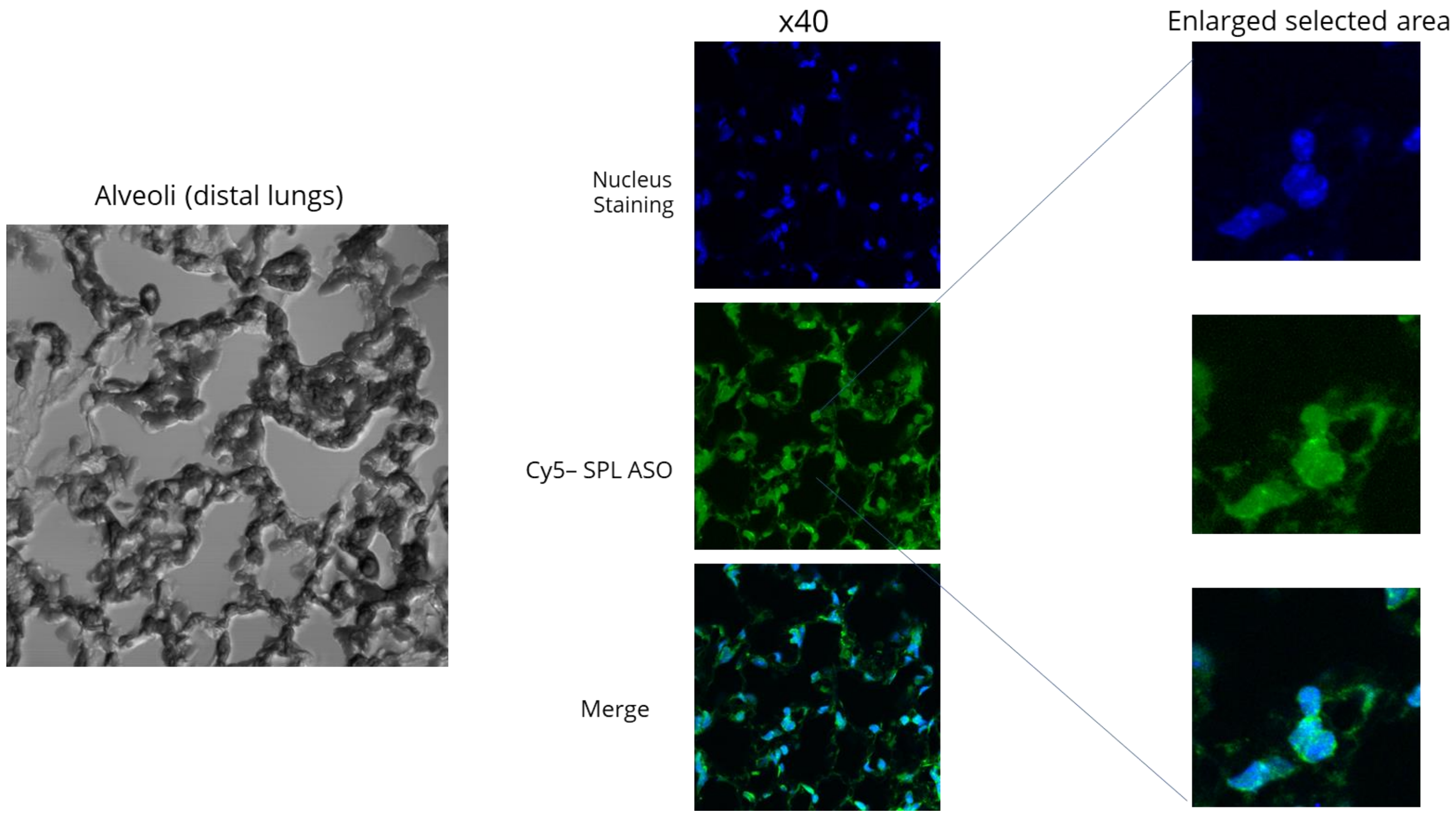


**SPL84 penetrates epithelial cell nuclei.** An example of a part of the alveoli from a mouse lung is presented. The presented confocal microscope image is a stack of only 4′,6-diamidino-2-phenylindole (DAPI) positive optical sections. Blue - nuclei (DAPI staining), Green - Cy5-SPL84.
